# An evaluation of the effectiveness of critical components of the chain of survival in out-of-hospital cardiopulmonary resuscitation in Nigeria

**DOI:** 10.1101/2024.05.31.24308305

**Authors:** Israel Kayode Kolawole, Lukman Olajide Abdur-Rahman, Shehu Abubakar Kana, Aisha Oluwabunmi Ajala, Akinyemi Aje, Olubusola Temitope Alagbe-Briggs, Yekeen Ayodele Ayoola, Nkeiruka Chigekwu Mbadiwe, Hadiza Shehu Galadanci, Josephus Kayode Ladipo, Dalhat Salahu, Arinola Adeyoola Sanusi, James Ayodele Ogunmodede, Tonia Chinyelu Onyeka, Tanimu Sambo Yusuf, Olaitan A Soyannwo, Kene Terfa

**Author notes:** **Corresponding Author**: Prof. Israel Kayode Kolawole, Department of Anaesthesia, Faculty of Clinical Sciences, College of Health Sciences, University of Ilorin, Ilorin, Nigeria.

## Abstract

**Background:** The ‘’Chain of Survival’’ (CoS), provides a structure to evaluate an emergency response system for out-of-hospital cardiac arrest (OHCA) in any community. This chain has not been previously evaluated in an African population. The aim of this study was to assess the effectiveness and efficiency of out-of-hospital CPR by evaluating the components of the CoS following sudden cardiac arrest in Nigeria.

**Methodology:** This was a prospective multicenter descriptive cross-sectional exploratory survey of epidemiology and outcome of sudden cardiac arrest (SCA) in adult Nigerians. The survey was conducted in six University teaching hospitals in the 6 geopolitical zones. Information concerning the patients were obtained from family member or an accompanying person, and hospital emergency department personnel. Exclusion criteria included age < 18 years, incomplete registered data, do not attempt resuscitation (DNAR) decision, and patients in whom resuscitation was not attempted. Primary outcome measures were return of spontaneous circulation (ROSC), survival to hospital admission and discharge.

**Results:** A total of 387 OHCA patients aged between 18 and 115 years met the inclusion criteria. They consisted of 62.5% males and 37.5% females, giving a male/female ratio of approximately 1.7:1, and the mean age was 49.9 ± 19.15 years. Most (55.6%) of the arrests occurred at home. Ninety-one-point seven percent (91.7%) of the arrests were witnessed. Calls were made to ambulance services but only in a third (34.6%) did ambulance arrive. The time taken for the ambulance to arrive at the call site ranged between 10 minutes and 60 minutes, with a mean time of arrival of 25.6 ±20.5 minutes. Only 14.3% of ambulances had facilities for both BLS interventions. Pre-hospital resuscitation was attempted in 15.2%. Pre-hospital ROSC was achieved in 3.6% of all the SCA events, but only 1.8% survived to hospital admission and were eventually discharged.

**Conclusion:** The emergency response service to OHCA in Nigeria is very poor due to weaknesses in the critical components of the CoS. There is a need for regular evaluation of the effectiveness of the CoS to identify weaknesses, and efforts made to improve performance in every component of the chain, through community wide BLS training.

**Clinical Perspective:** *What is new?:*

- This is the first large scale multi-center study in Nigeria, and probably in Africa, to evaluate the effectiveness and efficiency of the components of chain of survival in out-of-hospital cardiopulmonary resuscitation, reflecting the complex interplay of resources, setting and bystander engagement on the clinical outcomes of cardiac arrest.
- The emergency response service system for OHCA in Nigeria is very poor.
- This study revealed the shortcomings related to cardiopulmonary resuscitation due to resource-limitation, which bellies the practical difficulties inherent in implementing the scientific recommendations on resuscitation formulated primarily from the perspective of a high-resource environment by the International Liaison Committee on Resuscitation.

*Clinical implications:*

- The shortcomings observed in cardiopulmonary resuscitation in this study calls for a well-organized, coordinated deliberate efforts to strengthen all parts of the chain of survival in a manner that emphasizes engagement from the community, and the development of locally implemented, strong pre-hospital chain of care, which is adaptable to the requirements of different cardiac emergencies.
- There should be a long-term, ongoing evaluation of the CoS to identify weaknesses and efforts made to improve performance in every community through a deliberate effort to ensure training on BLS for everyone (medical and non-medical), and from an early age.

## INTRODUCTION

An out-of-hospital sudden cardiac arrest (OHCA) is defined as cessation of cardiac mechanical activity that occurs outside of the hospital setting and is confirmed by the absence of signs of circulation.^1^ It remains, a major public health challenge worldwide, with survival ranging between 2 and 11%.^2,3^

The term “Chain of Survival” describes a series of sequentially connected interdependent links of critical interventions designed to reduce the mortality associated with cardiac arrest. The concept was proposed in the 1992 CPR guidelines and provides a structure to evaluate an emergency response system for OHCA patients in any community for optimal outcome.^4^ The six structural components of the Chain include: early recognition and access to emergency medical care, early CPR, early defibrillation, early advanced cardiac life support (ACLS), integrated post-resuscitative care, and long-term rehabilitation. Strict adherence to the Chain is necessary for optimal outcome. However, international resuscitation guidelines are usually based on scientific recommendations, which are formulated typically from the perspective of well-resourced environment, with little consideration of applicability in LMIC.

This is the first large scale multi-center study in Nigeria, and probably in Africa, that have assessed the effectiveness and efficiency of the components of CoS in out-of-hospital CPR, reflecting the complex interplay of resources, setting and bystander engagement, on the clinical outcomes of cardiac arrest. The aim was to assess performance and identify gaps in the components of the Chain.

## METHODS

### Study setting

Nigeria is the most populous country in Africa, and the most populous black nation in the World,^5^ with an estimated population of about 206 million as at 2020.^6^ The country is a multi-ethnic, multi religious and multi-cultural society, officially subdivided into six geopolitical zones (Figure 1). This study was conducted in six purposefully selected University teaching hospitals in the six geopolitical zones. The hospitals are, University of Ilorin Teaching hospital (North Central), Aminu Kano teaching hospital (North-West), University College hospital, Ibadan (South-West), Gombe State University teaching hospital (North-East), University of Nigeria teaching hospital, Ituku-Ozalla, Enugu (South-East) and University of Port-Harcourt teaching hospital (South-South).

**Figure 1:**
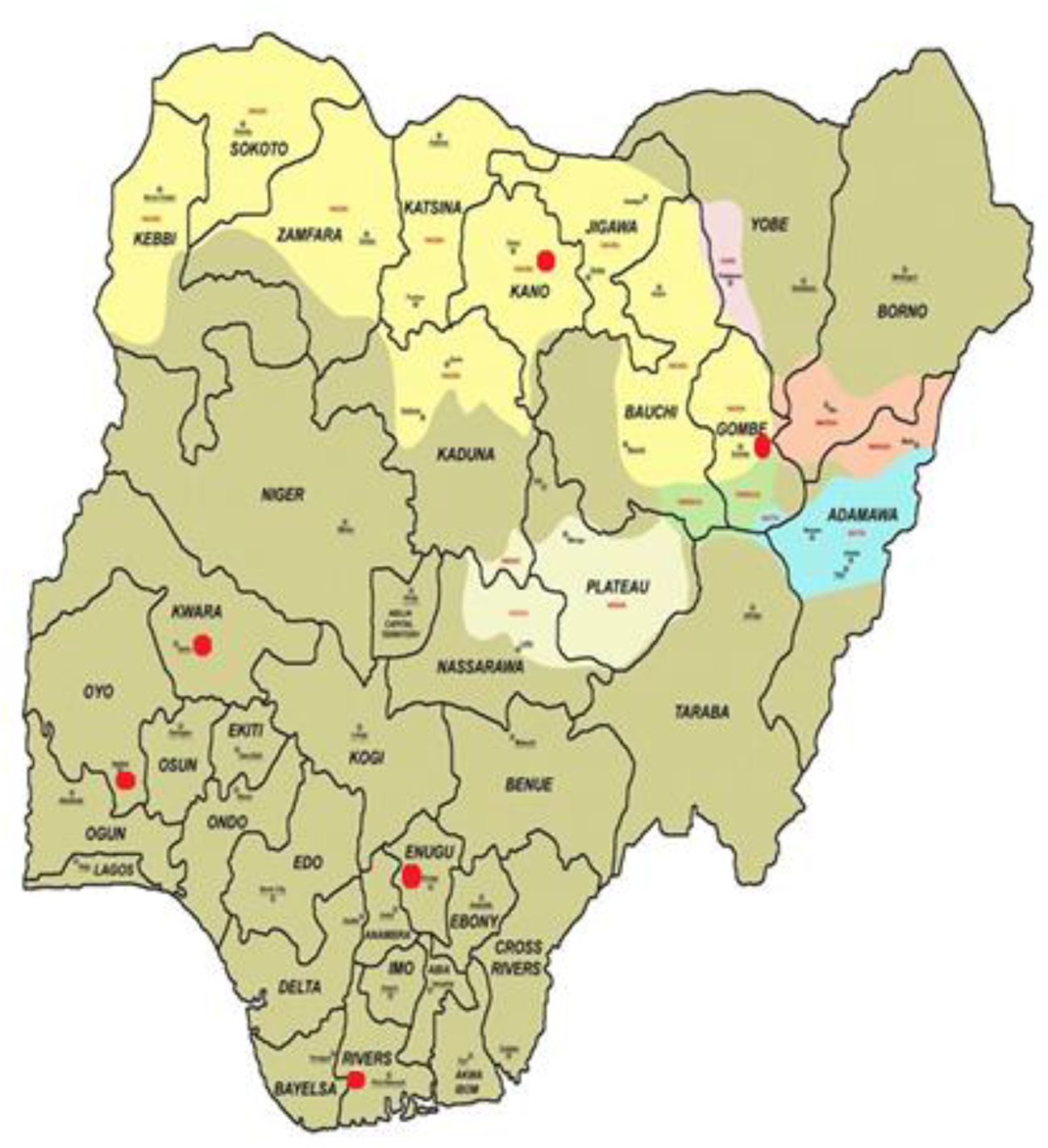
Map of Nigeria showing the _2_d_4_istribution of participating hospitals in the six geopolitical zones.

### Design

A sub-analysis of the data on OHCA extracted from a prospective multicenter descriptive cross-sectional exploratory survey of epidemiology and outcome of sudden cardiac arrest (SCA) in Nigerian adults was conducted.

### Participants

All adults (⩾ 18 years) who suffered OHCA and were transferred to the six selected teaching hospitals from 10^th^ November 2020 – 9^th^ July 2022 were recruited into the study.

### Exclusion criteria

Sudden cardiac arrest in patients that had non-cardiac chronic terminal illness like malignancy that was not in remission, incomplete data, or violation of protocol regarding age limits, and patients with ‘’do not attempt resuscitation’’ instructions (DNAR), and patients in whom resuscitation was not attempted were excluded.

### Main outcome measures

Main outcome measures were ROSC, survival to hospital admission and survival to hospital discharge.

### Compliance with Ethical Standards

The study was approved by the local ethics committee of each participating hospital and the National health research ethics committee (NHREC/01/01/2007-25/08/2020).

## STATISTICAL ANALYSIS

Statistical analysis was done using descriptive statistics (frequency, proportions, means and standard deviation) to summarize variables. Analysis of quantitative data was done using IBM-SPSS version 25.0 (SPSS Inc., Chicago, IL, USA. Data was appropriately presented using tables. Continuous variables were summarised using means and standard deviations for normally distributed variables, and any between group comparisons was tested with the student’s t-test or ANOVA as indicated. Categorical variables like gender, causes of SCA and treatment types were summarized as number (frequency tables) and percentages, and compared between groups with the Chi-square test and Yate corrected and Fishers’ exact tests as indicated. Survival was determined as the percentage of persons who survived to hospital discharge out of the number of persons with SCA. Bi-variate analysis (cross tabulation) was done to have an idea of how some factors relate directly to the outcome and the site of occurrence of the sudden cardiac arrest. All tests were considered significant at p<0.05.

## RESULTS

A total of 387 OHCA brought to the selected hospitals met the inclusion criteria for analysis. Table I shows the demographic and socio-cultural characteristics of the patients. There were 242 (62.5%) males and 145 (37.5) females, giving a male/female ratio of approximately 1.7:1. The age ranged between 18 years and 115 years, with a mean age of 49.9 ± 19.15), and median of 48.0 years. The patients were arbitrarily stratified into 3 age categories. The mean age of males was 49.9 ±19.35 years and the mean age of females was 49.8 ±18.86 years.

**Table I:** Demographic and Socio-Cultural Characteristics of the OHCA patients.

| **Parameters** | **Frequency**<br>**(N=387)** | **Percentage** |
| --- | --- | --- |
| **Patient Age** |  |  |
| <40 years | 126 | 32.6 |
| 40-64 Years | 167 | 43.1 |
| 65+ Years | 94 | 24.3 |
| **Patient gender** |  |  |
| Male | 242 | 62.5 |
| Female | 145 | 37.5 |
| **Marital status** |  |  |
| Married | 271 | 70.0 |
| Single | 76 | 19.6 |
| Widowed | 33 | 8.5 |
| Divorced/Separated | 7 | 1.8 |
| **Type of Residence** |  |  |
| Urban dwellers | 319 | 82.4 |
| Rural dwellers | 68 | 17.6 |
| **Living arrangements** |  |  |
| Lives with others | 370 | 95.6 |
| Lives alone | 17 | 4.4 |
| **Availability of Family Support** |  |  |
| Good: families around them all the time | 225 | 58.1 |
| Fair: families around them sometimes | 62 | 16.0 |
| Poor: don't have families around them | 14 | 3.6 |
| Unknown | 86 | 22.2 |
| **Socio-economic class** |  |  |
| 26-29: Upper (I) | 37 | 9.6 |
| 16-25: Upper Middle (II) | 182 | 47.0 |
| 11-15: Lower Middle (III) | 97 | 25.1 |
| 5-10: Upper Lower (IV) | 40 | 10.3 |
| Unknown | 31 | 8.0 |

Table II shows the location of arrests, witnesses to the arrest and responses to the patients. Most of the arrests 215 (55.6%) occurred at home, 147 (38.0%) occurred at public places, such as offices, market, roadside, party, place of worship and sports field, while 25 (6.5%) cases occurred in other places, (such as en-route to hospital, peripheral hospital, delivery home and others). Three hundred and fifty-five (91.7%) of the SCA were witnessed. Arrests were not witnessed in 32 (8.3%) of cases. A total of 351 (90.7%) patients possessed mobile phones. Calls were made to ambulance services in 26 (6.7%) of cases, but in only 9 (34.6%) of these cases did ambulance arrive, of which only 3 (42.9%) had facilities for providing life support interventions. The time taken for the ambulance service to arrive to the call site ranged between 10 minutes and 60 minutes with a mean time of arrival of 25.6 ±20.5 minutes (median =10 minutes). Fifty-five-point six percent of the ambulances arrived between 10 and 19 minutes, while the remaining 44.4% arrived after 29 minutes. Ambulance did not arrive at all in 17 (65.4%) of the cases, while calls were not made for ambulance services in 361 (93.3%) of the cases. Private or public transportation was the available means of conveying the patients to hospital in 369 (95.3%) of cases, while ambulances were used in 7 (1.8%) of cases, 4 (57.1%) of which had no means of providing life support interventions. Two (28.6%) of the ambulances had facilities for both basic-life support (BLS) interventions and life-saving drugs without a trained health personnel and only one (14.3%) of the ambulances was equipped with facilities for BLS intervention, life-saving drugs, defibrillators, and personnel trained to deal with emergencies. The mode of pre-hospital transportation was not specified in 11 (2.8%) of the cases.

**Table II:** Location of arrest, Witness to the arrest, Possession of mobile phones, activation of Emergency Medical System and means of transportation of patients to health facilities.

| Parameters | Frequency (N =387) | Percentage |
| --- | --- | --- |
| **Location of arrest** |  |  |
| Home | 215 | 55.6 |
| Public places – Office, Market, Roadside, Party, Place of worship, Sports field, Hotel room | 147 | 38.0 |
| En-route to hospital | 11 | 2.8 |
| Peripheral hospital | 8 | 2.1 |
| Delivery home | 4 | 1.0 |
| Unspecified | 2 | 0.5 |
| **Arrest witnessed or not witnessed** |  |  |
| Arrest witnessed | 355 | 91.7 |
| Arrest not witnessed | 32 | 8.3 |
| **Who witnessed?** |  |  |
| Family bystanders | 242 | 62.5 |
| Public bystanders | 87 | 22.5 |
| Health personnels | 26 | 6.7 |
| **Possession of mobile phone (N =387)** |  |  |
| Yes | 351 | 90.7 |
| No | 36 | 9.3 |
| **Activation of Emergency Medical System** |  |  |
| **Was call made for ambulance service?** |  |  |
| No | 361 | 93.3 |
| Yes | 26 | 6.7 |
| **Did any ambulance arrive? (N =26)** |  |  |
| No | 17 | 65.4 |
| Yes | 9 | 34.6 |
| **Facilities inside the ambulances (N =7)** |  |  |
| Ambulances without life-support interventions | 4 | 57.1 |
| Ambulances with facilities for both BLS and life-saving drugs, without a trained health personnel | 2 | 28.6 |
| Ambulances with facilities for BLS, life-saving drugs, defibrillators, and trained personnel | 1 | 14.3 |
| **Estimated Time of ambulance arrival** |  |  |
| 10-19 minutes | 5 | 55.6 |
| 20-29 | 0 | - |
| 30-39 minutes | 2 | 22.2 |
| 40-49 minutes | 0 | - |
| ≥50 minutes | 2 | 22.2 |
| Mean time (minutes) | 25.6 ±20.5 | Median =10 |
| **Type of pre-hospital transportation (N =387)** |  |  |
| Private or public transportation | 369 | 95.3 |
| Ambulances | 7 | 1.8 |
| Unspecified | 11 | 2.8 |

Table III shows the pre-hospital medical interventions. Pre-hospital intervention (resuscitation) was attempted, either at the scene or during transportation to health facilities, in 59 (15.2%) cases. The first monitored rhythms in the OHCA were pulseless VT in 2 (3.4%) victims, asystole in 1 (1.7%) and bradycardia in another (1.7%). The rhythm status was not monitored for 55 (93.2%) of the cases.

**Table III:** Pre-hospital medical intervention.

| Parameters | Frequency | Percentage (%) |
| --- | --- | --- |
| **Resuscitation (N=387)** |  |  |
| Attempted | 59 | 15.2 |
| Not attempted | 328 | 84.8 |
| **Reasons for resuscitation not attempted (n =328)** |  |  |
| Considered futile | 134 | 34.6 |
| Brought in dead | 10 | 2.6 |
| Others | 20 | 5.2 |
| Unknown | 164 | 42.4 |
| **Who instituted resuscitation? (N =59)** |  |  |
| Family Bystanders | 25 | 42.4 |
| Public Bystanders | 19 | 32.2 |
| Trained health personnel | 15 | 25.4 |
| **Number of people available to institute CPR (N =59)** |  |  |
| Multiple rescuer | 30 | 50.8 |
| Two rescuers | 16 | 27.1 |
| Single rescuer | 13 | 22.0 |
| **Equipment used during resuscitation (N=59)** |  |  |
| Bag-mask device (AMBU Bag) | 29 | 49.2 |
| Airway devices: | 12 | 20.3 |
| Oropharyngeal/Nasopharyngeal/Laryngeal mask airway/Tracheal Tube |  |  |
| ECG monitor/AED/Manual defibrillator | 2 | 3.4 |
| Barrier device – Pocket mask | 2 | 3.4 |
| Unspecified | 14 | 23.7 |
| **First monitored rhythm** |  |  |
| Pulse less VT | 2 | 3.4 |
| Asystole | 1 | 1.7 |
| Bradycardia | 1 | 1.7 |
| Unknown | 55 | 93.2 |

Table IV and V show the age and gender distribution of the patients, location of arrest, witnesses to arrest, who instituted resuscitation, place of post cardiac arrest care and outcomes of resuscitation. Pre-hospital ROSC was achieved in 14 (23.7%) of 59 resuscitation attempts, but only 7 (11.9%) patients, accounting for (50.0%) of the pre-hospital ROSC survived to hospital admission and eventually survived to hospital discharge, with an overall hospital discharge survival of 7 out of 387 SCA (1.8%). The remaining 7 (50.0%) pre-hospital ROSC suffered re-arrest en-route hospital. Pre-hospital ROSC and survival to hospital admission and discharge were significantly better in the females than males, (p < 0.05). Of all the 147 (38.0%) cases of SCA that occurred in the public places, only 23 (15.6%) benefited from bystander CPR interventions, which yielded 4 (17.4%) ROSC, none of whom survived to hospital admission. Resuscitation was attempted in 24 (11.2%) of the 215 patients who had SCA at home, out of which only 3 (12.5%) achieved ROSC, but only 1 survived to hospital admission and was eventually discharged from hospital alive.

**Table IV:** Age and gender distribution of OHCA patients, location of arrest, witnesses to arrest and who instituted resuscitation VS outcome.

| Parameters | Frequency/Percenta | Pre-hospital | Survival to | Survival to |
| --- | --- | --- | --- | --- |

|  | ge (N =387) | ROSC n =14<br>(3.6%) | hospital admission<br>n =7 (1.8%) | hospital discharge<br>n =7 (1.8%) |
| --- | --- | --- | --- | --- |
| **Age in Years** |  |  |  |  |
| < 65 years (N =50 or<br>84.7%) | 293(75.7) | 11 (22.0) | 5 (10.0) | 5 (10.0) |
| ≥ 65 Years (N=9 or<br>15.3%) | 94 (24.3) | 3 (33.3) | 2 (22.2) | 2 (22.2) |
| **p Value** | Not Significant | 0.462 | 0.296 | 0.296 |
| **Gender** |  |  |  |  |
| Male | 242 (62.5) | 4 (1.7) | 1 (0.4) | 1 (0.4) |
| Female | 145 (37.5) | 10 (6.9) | 6 (4.1) | 6 (4.1) |
| **p Value** | Statistically Significant | 0.007 | 0.023<sup>y</sup> | 0.023<sup>y</sup> |
| **Location of arrest** |  |  |  |  |
| Home N=215<br>(55.6%) | 24 (11.2) | 3 (12.5) | 1 (4.2) | 1 (4.2) |
| Public places<br>N=147(38.0%) | 23 (15.6) | 4 (17.4) | 0 | 0 |
| Other places N=25<br>(6.5%) | 12 (48.0) | 7 (58.3) | 6 (50.0) | 6 (50.0) |
| **p Value** | Statistically Significant | 0.006 | 0.001<sup>y</sup> | 0.001<sup>y</sup> |
| **Witnesses to arrest**<br>**(N =355)** |  |  |  |  |
| Family members (N<br>=242) | 242 (68.2) | 4 (1.7) | 1 (0.4) | 1 (0.4) |
| Public bystanders (N<br>=87) | 87 (24.5) | 3 (3.4) | 0 (0.0) | 0 (0.0) |
| Trained health<br>personnel (N =26) | 26 (7.3) | 7 (26.9) | 6 (23.0) | 6 (23.0) |
| **p Value** | Statistically Significant | 0.001 | 0.001<sup>y</sup> | 0.001<sup>y</sup> |
| **Who instituted**<br>**resuscitation (N =59)** |  |  |  |  |
| Bystanders (N =25<br>Family members + 19<br>Public =44) | 44 (74.6) | 7(15.9) | 1 (2.3) | 1 (2.3) |
| Trained health<br>personnel (N =15) | 15 (25.4) | 7 (46.7) | 6 (40.0) | 6 (40.0) |
| **p Value** | Statistically Significant | 0.020 | 0.001 | 0.001 |

<sup>\*y</sup> =Yates correction

**Table V:** Outcome of resuscitation Outcome measures.

| Outcome measures | Frequency | Percentage |
| --- | --- | --- |
| **Pre-hospital return of spontaneous circulation (ROSC) (N** |  |  |
| =59) | 14 | 23.7 |
| Yes |  |  |
| No | 45 | 76.3 |
| **Survival to hospital admission (N =14)** |  |  |
| Survived | 7 | 50.0 |
| Suffered re-arrest and died during transport to hospital | 7 | 50.0 |
| **Place of Post Arrest Care in the hospital (N =7)** |  |  |
| ICU | 4 | 57.1 |
| HDU | 1 | 14.3 |
| Open ward | 2 | 28.6 |
| **Overall Hospital discharge survival (N= 387)** |  |  |
| Survived to discharge | 7 | 1.8 |
| **Neurological status at discharge (N =7)** |  |  |
| CPC 1 or 2 | 4 | 57.1 |
| CPC 3 | 3 | 42.9 |

\*CPC =Cerebral Performance Category

Resuscitation was attempted in 12 (48.0%) of the 25 patients who had SCA in other places, apart from home and public places, (such as en-route to hospital, peripheral hospital, delivery home and others), out of which 7 (58.3%) achieved ROSC, and 6 (50.0%) survived to hospital admission and were eventually discharged from hospital alive. Proportionately, 12 (48.0%) of the arrests that occurred in places such as en-route to hospital, peripheral hospital and delivery homes benefited from pre-hospital intervention compared to those who arrested in other places. Survival was also significantly better in terms of pre-hospital ROSC, survival to hospital admission and survival to eventual discharge from hospital in this category of patients (p =0.0001) compared to the other categories. Majority, 44 of 59 (74.6%) of the pre-hospital CPR intervention were provided by bystanders, while only 15 (25.4) were provided by trained health personnel. However, most (7 or 50.0%) of the pre-hospital ROSC were from patients resuscitated by healthcare personnel and 6 (85.7%) of them survived to hospital admission and were eventually discharged from hospital alive. This is unlike the 7 (50.0%) pre-hospital ROSC resuscitated by bystanders, out of which only 1 (14.3%) survived to hospital admission and was eventually discharged from the hospital alive. These differences were statistically significant for ROSC (p <0.05). The ROSC, survival to hospital admission and discharge were also significantly more in the CPR provided by the trained health personnel (p < 0.05).

Post cardiac arrest hospital care was done for 4 (57.1%) of the 7 admitted patients with ROSC in the intensive care unit (ICU), 1 (14.3%) in the high dependency unit (HDU) and 2 (28.6%) in the open ward. All the patients (100.0%) survived to discharge regardless of the area of post arrest care. The neurological outcome at discharge, assessed using the Cerebral Performance Category (CPC) scale, showed that 4 (57.1%) of the survivors were discharged with a favorable neurological outcome (CPC of 1 or 2), and 3 (42.9%) with unfavorable neurological outcome (CPC of 3).

## DISCUSSION

In this study, we used three working definitions of survival as described in previous studies.^7,4^ These include (1) return of spontaneous circulation (ROSC), (2) successful hospital admission, defined as sustained ROSC till hospital admission and (3) successful hospital discharge, defined as the patient leaving the hospital alive. However, the survival rate to hospital discharge is regarded as the “gold standard” for determining the effectiveness of treatment of cardiac arrest.^4^

The overall outcomes from OHCA in this study are poor both in terms of survival, and neurologic and functional ability. The rate of pre-hospital CPR intervention (15.2%) was low. Despite the 23.7% of ROSC achieved from the pre-hospital CPR intervention, the overall survival to hospital admission and eventual discharge from hospital was only 1.8%. This finding is abysmal compared to the reports from developed countries.^8,9^ This is indicative of the poor quality of pre-hospital CPR intervention and emergency care in our setting. The morbidity and mortality weekly report of the Cardiac Arrest Registry to Enhance Survival (CARES) – an out-of-hospital cardiac arrest surveillance, recorded an overall survival to hospital discharge rate of 9.6%,^1^ which according to the authors was higher than in some previous reports.^10^ Reports put the median rates of survival to hospital discharge from OHCA at 7.9%.^11,10^ However, in interpreting and comparing the findings of this study with any previous report from another population, we need to consider the context and the standard of care available for out-of-hospital medical emergencies in the different populations. Survival from OHCA varies widely across countries and geographical regions, depending on the characteristics of emergency medical services (EMS) and health care systems in the community, such as treatment availability, training, and care quality.^8^ Reported outcomes of OHCA in low-resource settings are usually lower than those reported in patients from high-resource countries, obviously due to fewer available resources for resuscitation care compared to high-resource regions.^3^ According to the World Health Organization, ‘‘the state of cardiac arrest and trauma resuscitation is a good reflection of the standard of pre-hospital and emergency care in any health system.”^12^ Our findings, therefore, reflect the weaknesses in community emergency medical care service system in Nigeria compared to developed world.

Nigeria, by the World Bank classification, is a lower middle-income country. The healthcare service provision is cash and carry, based on out-of-pocket payments. Consequently, access to healthcare, including emergencies, remains a very big challenge, severely limited by ability of the citizens to pay for care cost. The country faces major challenges in each of the six areas in the links of the CoS in terms of timing, effectiveness, and efficiency of execution of the components of the chain. There was poor ambulance response time and more importantly, low level of community participation as shown by poor bystander CPR intervention and lack of trained EMS personnel. This is due to lack of organized pre-hospital emergency medical services, and basic infrastructural deficits. In fact, as clearly shown from the findings of this study, it is obvious that the concept of chain of survival is not well rooted in the out-of-hospital emergency medical services in Nigeria.

Early access, which is the first link in the Chain of Survival, is regarded as the most essential component of the chain.^13^ However, where the arrest is not witnessed, as happened in 8.3% of cases in this study, the link will obviously be compromised. Previous study reported survival from unwitnessed cardiac arrest to be only 4% compared with 22% for witnessed arrests.^14^ There was no record of CPR attempt in cases of unwitnessed arrest in our study.

Incidentally, majority (91.7%) of the OHCA events in this study were witnessed. This volume of witnesses is large compared to what was reported in a previous study in South Africa (68%),^15^ and in a morbidity and mortality weekly report of CARES in the United States,^1^ and in a similar study in Canada.^16^

The benefit of an arrest being witnessed is the activation of emergency response system. The large volume of witnessed arrest in our study was, therefore, expected to facilitate the activation of the emergency medical system, which is an important part of early access.

However, this is only possible where there is a dedicated nationally promoted and well publicized emergency telephone number/code for such communication. Unfortunately, providing toll free emergency response services through a dedicated unique telephone number, like 911 in the US, is a herculean task and almost an impossibility in most communities in Nigeria. This is because, a look at the displays of the emergency telephone number used in many countries throughout the world did not show Nigeria on the list.^17^ An area-wide emergency toll-free telephone numbers/code, as obtains in the UK and North America, is either non-existent, or if at all exists in Nigeria, is not well publicized. Enhancing emergency medical response service through a dedicated unique telephone number in all communities should therefore be a top priority for Nigeria. Widespread use of dedicated emergency telephone numbers has simplified and shortened access to emergency assistance in most advanced/developed communities.

The home location of the arrests in this study ensured that 68.2% of the witnesses were family members (family bystanders). However, the advantage of this to the survival of the patients, in the form of early access to ensure prompt intervention, was lost in this study in the absence of prompt notification of the cardiac arrest response team, and an inefficient out-of-hospital emergency medical evacuation system (EMS). Nonetheless, the finding reinforces the need to include family members in the education efforts and information dissemination to patients at risk of sudden death in order to ensure timely and effective delivery of interventions by bystanders. The various categories of arrest witnesses in this study may also have contributed to the telephone calls made for ambulance services. Unfortunately, despite the widespread claim of good mobile telephone penetration within the communities in Nigeria, as attested to in this study with 90.7% of the victims possessing mobile phones, calls could only be made for ambulance services in 6.7% of cases.

The available emergency medical care system, which can be activated for OHCA in Nigeria are essentially hospital-based ambulance services. These out-of-hospital ambulance services are operated by both public and private hospitals. Furthermore, the ambulance service system in Nigeria is inefficient and bedeviled with numerous structural challenges, such as delayed response time. The fact of this situation was buttressed by the findings of this study in which majority (55.6%) of the ambulances in this study arrived much late; arriving between 10 and 19 minutes, while a sizable number (44.4%) arrived after 29 minutes. The median time of ambulance arrival was 10 minutes. This is a period long enough for permanent brain damage to ensue in the absence of any effective CPR intervention by the bystanders. A study in South Africa, which is classified as an upper middle-income country by the World Bank, found that the median response time for OHCA cases was 9 min.^15^ Many of the ambulances in Nigeria are mere conveyor or evacuation vehicles and not necessarily designed for life-saving interventions. They are often too poorly equipped and lacked appropriately trained personnel and facilities to provide BLS and AED interventions required for community emergency medical services. A good ambulance services must have adequate facilities for emergency medical intervention. At the least, the service according to the European Resuscitation Council guidelines, must have a driver with a paramedical background and a nurse, both trained and qualified to perform Advanced Cardiac Life Support (ACLS), with expertise to render such services as advanced airway interventions, administration of inotropic drugs and use of a defibrillator.^18^

The fact that ambulance transportation occurred in only 77.8% of the 9 ambulance responses suggest that, even some of the very few ambulances that responded to calls probably arrived after the patients had been transported to hospital in private or commercial vehicles. In most cases, transportation of patients to health facilities for emergency medical intervention is in Nigeria done through private arrangements using any available vehicle from family, friends or commercial taxis. This is attested to by the findings of this study, in which a high majority (95.3%) of the patients were transported to the hospital by private or public transportation.

These vehicles were unlikely to have facilities for medical interventions since they were not designed for such purpose, and the survival of such patients to the hospital cannot be guaranteed. The fact that 50% of patients with pre-hospital ROSC died en-route to hospital before hospital admission demonstrate inadequate facilities / or knowledge and skills of BLS in the ambulance personnel or the accompanying bystanders in the alternative means of transportation used to convey the patients to hospital. In other words, the resources available within the community (ambulance facilities/equipment and personnel) to convey the patients to hospital could not sustain the gain already made at the scene of the event.

Bystander CPR is the second link in the CoS and represents the only chance for a victim of sudden cardiac arrest to survive.^19^ The time to initiation of bystander CPR, (the time from collapse to initiation of CPR), could not be ascertained in this study because it was not monitored. It has been demonstrated that the probability of survival approximately doubles when bystanders initiate CPR before the arrival of EMS personnel.^20^ However, the results of the present study show that, the achievement of anything close to this is hampered by a number of challenges in Nigeria due to poorly organized community emergency medical response system, poor bystander intervention and inadequate resources in terms of personnel and facilities to mount a timely and effective intervention.

The fact that only 21.8% of public bystanders witnessed arrests benefited from BLS intervention in this study indicates poor knowledge / or willingness to perform this intervention within the community. The rates of bystander CPR remain poor in most low-middle income countries (LMIC).^21^ This reflects the level of community involvement in the treatment of OHCA and forms an important modifiable factor in any efforts to improve the survival of OHCA. It highlights the importance of deliberate, coordinated, and effective community-based CPR training with efforts made to teach community members about the different ways in which out-of-hospital cardiac arrest patients can present, and the need to initiate CPR, and use AEDsin the event of SCA. Countries across the world have tried to address the issue of low bystander intervention within the community by introducing mandatory CPR training at various opportunities, levels and strata of their society.^22^ The potency of this is verifiable with the experience of Norway in which introduction of CPR training into the national school education curriculum since 1961, has succeeded in raising the average survival from OHCA to about 25%.^23^

The fear of getting involved because of the distrust of law enforcement, as identified in a previous study in minority low-income neighbourhood community in the United State of America,^24^ is also relevant and constitute a huge potential barrier to willingness to perform bystander CPR in the LMIC, including Nigeria. To improve bystander action in cases of OHCA in Nigeria therefore, there is need to put in place appropriate laws to absolve the bystander who provides reasonable aid/offer assistance to victims of SCA in the community for the victim’s health condition and protect him/her from prosecution if anything goes wrong. Such laws will encourage laypersons to willingly help without any hesitation.

Although the CPR knowledge and skill of the family members and bystanders who witnessed the arrest in our study could not be ascertained, the findings of the study demonstrate a poor knowledge and skills of BLS within the community. This is attested to by the fact that, even with the 15.2% pre-hospital CPR interventions, only a small percentage (23.7%) of pre-hospital ROSC was achieved, with an eventual hospital admission and discharge survival of 11.9% respectively. The 1.8% overall hospital discharge survival out of the total 387 OHCA events analyzed for the study is abysmally low compared to the overall survival rate of 11.2% reported in the CARES study,^1^ and the average survival of 25% in Norway.^23^ This shows that even when an arrest is witnessed, the success of CPR, if at all instituted, will depend on the speed of initiation and the knowledge and skills of the operator. Deficient resuscitation skill within the community is a risk factor responsible for poor outcome after out-of-hospital cardiac arrest, even in witnessed arrests. However, it is good enough that some of the bystanders, (family bystanders and public bystanders), initiated the first link in the chain of survival by providing BLS interventions to some of the patients. The efforts of the bystanders (family and public) in this study yielded an overall 15.9% of pre-hospital ROSC in all the cases of their 74.6% CPR interventions. On the other hand, the 25.4% of the trained health personnel-initiated CPR achieved 46.7% pre-hospital ROSC. This clearly shows the benefits of knowledge and skills in achieving good outcomes in CPR. These healthcare workers, apart from participating in the initial resuscitation efforts, may also have assisted in arranging prompt transportation of the patient to hospital.

Our study revealed that, the rate of bystander-initiated CPR was substantially lower (10.3%) if the OHCA happened in the patient’s home compared with arrest in the public places (21.8%). In fact, the proportion of BLS intervention (21.8%) in public bystander witnessed arrest was twice the proportion of BLS intervention (10.3%) in family bystander witnessed arrest. The low family bystander CPR is similar to the findings of previous studies.^25,26^ However, contrary to their explanation that the lower rate of bystander CPR at home might in some degree be explained by patients living alone, a large proportion of our patients (95.6%) live with others, which is a consequent of communal living arrangement prevalent in our communities. The only plausible reason that could be adduced to lower rate of bystander CPR at home, therefore, would be the fact that the spouses/relatives/living partners were less disposed to initiating CPR, either due to inadequate awareness, fear, or lack of skills. Reason such as this has been documented previously.^24,26^

The third link in the Chain of Survival is early defibrillation. Although, generally, each link in the CoS is important and the relative importance of each of the links has not been clearly demonstrated, rapid defibrillation is often considered to be the single most important factor in determining survival from adult sudden cardiac arrest. Investigators have observed that it is the interaction of early CPR with early defibrillation that is the most powerful intervention for ensuring optimal patient survival.^4^ Vaillancourt et al,^16^ identified bystander CPR and early first responder defibrillation as significant contributors to increased survival in OHCA. This is because only defibrillation can reverse a ventricular fibrillation (VF) arrest. Defibrillation is often provided in OHCA by bystanders or trained BLS first responders, if a defibrillator is available, prior to the arrival of EMS providers. However, as revealed in this study, availability of an AED is a rarity for emergency medical response in public places within the community in Nigeria. The ambulances hardly have the necessary facilities and personnel with requisite knowledge to provide emergency medical interventions. Many are hardly equipped with automated defibrillators or never accompanied by paramedics, physicians, or nurses to provide BLS-defibrillation. Of the 9 ambulances that responded for emergency calls, only 3 had life-support interventions. Although one had a defibrillator the ambulance personnel lacked sufficient training to make use of the AED.

The initial documented cardiac rhythm at the time of the arrest has been found to be an important determinant of survival, with the VF or VT constituting the bulk of the patients in whom CPR is beneficial following SCA.^27,28^ In a previous study by Tresch et al,^28^ all the patients who demonstrated VF as the initial out-of-hospital rhythm had the highest immediate successful resuscitation and hospital survival rates with approximately 43% successfully resuscitated and 21% surviving. On the other hand, patients who demonstrated asystole or electromechanical dissociation had the lowest successful resuscitation and survival rates.

Unfortunately, rhythm identification was not possible in the large majority of patients in this study, except in (6.8%) of the 59 resuscitation attempts due to lack of resources (personnel and materials) for such within the community. Of these cases in which rhythm identification was possible, only 2 patients (3.4%) had rhythms considered to be shockable (VF), which may explain the low survival rate in our study population. The rhythms in the remaining 2 patients, (1.7% each of bradycardia and asystole respectively), were not shockable and have usually been associated with poor prognosis, though some reports have recorded successful treatment with survival to hospital discharge even in asystole cardiac arrests.^29,30^

The fourth critical link in the chain for management of cardiac arrest is early effective ACLS (intubation, intravenous access and intravenous medications), which is commonly provided by paramedics, physicians or nurses at the scene or in-transit. As revealed by the findings of our survey, the present ambulance service system in Nigeria can hardly meet this level of infrastructure, facilities and personnel, which explains why the outcome is so poor. There were no records of the use of definitive airway management in our study, however, intravenous medications were administered in very few cases, suggesting that some patients had intravenous access established for drug administration.

The fifth link in the CoS is integrated post-resuscitative care. A person who survives SCA is usually admitted to the hospital for observation and treatment where the heart is monitored closely, medicines are given to reduce the chance of another SCA, and tests are performed to identify the cause of the SCA. The level of care received will depend on the facilities available in the different areas of the hospital. The 2010 ACLS guidelines recommend that all post-cardiac arrest patients should be managed in an ICU.^31^ As revealed in this study, some of the post-cardiac arrest patients were managed in other hospital locations apart from the ICU. This may be as a result of limited ICU bed capacity in some of the study sites.

However, it appears that the Nigeria healthcare system is not doing badly in this link of the chain of survival, as all the ROSC delivered to the hospital survived till discharge regardless of the area of the hospital where they received the post cardiac arrest care.

Good neurological outcome is generally defined as a Cerebral Performance Category (CPC) ≤ 2 or a modified Ranking scale (mRS) score of ≤3; both broadly representing the ability to live independently.^32^ The neurological outcome at discharge in this study, with just 1.0% of all the SCA events achieving a favorable CPC of ≤ 2 and 0.8% CPC of 3, is very poor. This is probably due to the various interplay of a complex of factors, such as delayed initiation of pre-hospital CPR, quality of CPR/defibrillation and unfavorable rhythms of the arrests, resulting in prolonged no flow and low flow phases of the arrests, with significant degrees of brain damage.

### Limitations

There were several limitations of this study. First, the study is limited by the fact that it was a descriptive cross-sectional exploratory survey and conducted only in some hospitals that were purposefully selected for representation of the geopolitical zones. This could have introduced some degree of selection bias. Secondly, several time-related events in the chain of survival that may be associated with survival from cardiac arrest were not specifically captured in this study. These time-related events include access time, (the time from collapse to the summoning of emergency aid), time to initiation of cardiopulmonary resuscitation (CPR), (the time from collapse to initiation of CPR) and time to definitive care, (the time from collapse to provision of defibrillation, intubation, or emergency medication). Thus, the prognostic significance of these time-related events could not be tested in this study.

### Conclusion

Despite the various limitations to the study, the strengths of our study lie in the fact that it was a prospective multicenter survey, conducted in tertiary referral hospitals providing inpatient and outpatient services, as well as emergency and critical care services in the six geopolitical zones of the country. The study revealed the shortcomings related to cardiopulmonary resuscitation due to resource-limitation in a low-resource setting like Nigeria. This bellies the practical difficulties inherent in implementing the scientific recommendations on resuscitation formulated primarily from the perspective of a high-resource environment by the International Liaison Committee on Resuscitation (ILCOR).

## Data Availability

The data are available

## Non-standard Abbreviation

CoS: Chain of survival

## Acknowledgements

We are immensely indebted to late Emeritus Professor Nimi Briggs for his assistance at every stage of the project. We also acknowledge the assistance of the West African College of Surgeons (WACS) and the Endowment Fund Committee (Nigeria) of WACS for the encouragement, moral, technical and logistic support at every stage of the project.

## Source of Funding

The projeact was supported by a grant obtained from the Tertiary Education Trust Fund (TETFund) of the Federal Republic of Nigeria (Grant number TETFUND/DR&D-CE/NRF/2019, BATCH 6).

## Notes

### Competing Interest Statement

The authors have declared no competing interest.

### Funding Statement

The project was supported by a grant obtained from the Tertiary Education Trust Fund (TETFund) of the Federal Republic of Nigeria (Grant number TETFUND/DR&D-CE/NRF/2019, BATCH 6).

### Author Declarations

The study was approved by the National health research ethics committee (NHREC/01/01/2007-25/08/2020)

## References

1. Frieden TR, Harold Jaffe DW, Thacker SB, Moolenaar RL, LaPete MA, Martinroe JC, et al. Morbidity and Mortality Weekly Report Out-of-Hospital Cardiac Arrest Surveillance-Cardiac Arrest Registry to Enhance Survival (CARES), Centers for Disease Control and Prevention MMWR Editorial and Production Staff MMWR Editorial Board. 2011;

2. Yan S, Gan Y, Jiang N, Wang R, Chen Y, Luo Z, et al. The global survival rate among adult out-of-hospital cardiac arrest patients who received cardiopulmonary resuscitation: A systematic review and meta-analysis. Crit Care. BioMed Central Ltd.; 2020 Feb 22;24(1):1–13.

3. Berdowski J, Berg RA, Tijssen JGP, Koster RW. Global incidences of out-of-hospital cardiac arrest and survival rates: Systematic review of 67 prospective studies. Resuscitation [Internet]. Elsevier Ireland Ltd; 2010;81(11):1479–87. Available from: 10.1016/j.resuscitation.2010.08.006

4. 4. Guidelines 2000 for Cardiopulmonary Resuscitation and Emergency Cardiovascular Care. Part 12: from science to survival: strengthening the chain of survival in every community. The American Heart Association in collaboration with the International Liaison Committee on Resuscitation - PubMed [Internet]. [cited 2023 Jan 26]. Available from: https://pubmed.ncbi.nlm.nih.gov/10966681/

5. Population in Africa, by country 2020 | Statista [Internet]. [cited 2023 Jan 26]. Available from: https://www.statista.com/statistics/1121246/population-in-africa-by-country/

6. Nigeria Population (2023) - Worldometer [Internet]. [cited 2023 Mar 4]. Available from: https://www.worldometers.info/world-population/nigeria-population/

7. Eisenberg MS, Cummins RO, Damon S, Larsen MP, Hearne TR. Survival rates from out-of-hospital cardiac arrest: recommendations for uniform definitions and data to report. Ann Emerg Med. Ann Emerg Med; 1990;19(11):1249–59.

8. Institute of Medicine of the National Academies. Strategies to Improve Cardiac Arrest Survival A Time to Act. Report Brief. 2015.

9. Vellano K, Crouch A, Rajdev M, Mcnally B. Cardiac Arrest Registry to Enhance Survival (CARES) Report on the Public Health Burden of Out-of-Hospital Cardiac Arrest Institute of Medicine. 2015;

10. Sasson C, Rogers MAM, Dahl J, Kellermann AL. Predictors of survival from out-of-hospital cardiac arrest: a systematic review and meta-analysis. Circ Cardiovasc Qual Outcomes. Circ Cardiovasc Qual Outcomes; 2010 Jan;3(1):63–81.

11. Nichol G, Thomas E, Callaway CW, Hedges J, Powell JL, Aufderheide TP, et al. REGIONAL VARIATION IN OUT-OF-HOSPITAL CARDIAC ARREST INCIDENCE AND OUTCOME. JAMA : the journal of the American Medical Association. NIH Public Access; 2008 Sep 9;300(12):1423.

12. Aufderheide TP, Nolan JP, Jacobs IG, Van Belle G, Bobrow BJ, Marshall J, et al. Global health and emergency care: a resuscitation research agenda–part 1. Acad Emerg Med. Acad Emerg Med; 2013 Dec;20(12):1289–96.

13. Cummins R.O, Chamberlain D.A, Abramson N.S et al. Recommended guidelines for uniform reporting of data from out-of-hospital cardiac arrest: the Utstein Style. A statement for health professionals from a task force of the American Heart Association, the European Resuscitation Council, the Heart and Stroke. Circulation. 1991;84(2):960–75.

14. Eisenberg MS, Bergner L, Hallstrom A. Cardiac Resuscitation in the Community: Importance of Rapid Provision and Implications for Program Planning. JAMA. American Medical Association; 1979 May 4;241(18):1905–7.

15. Stein C. Out-of-hospital cardiac arrest cases in Johannesburg, South Africa: A first glimpse of short-term outcomes from a paramedic clinical learning database. Emergency Medicine Journal [Internet]. 2009 Sep [cited 2023 Jun 27];26 (9):670–4. Available from: https://www.researchgate.net/publication/26760936_Out-of-hospital_cardiac_arrest_cases_in_Johannesburg_South_Africa_A_first_glimpse_of_short-term_outcomes_from_a_paramedic_clinical_learning_database

16. Vaillancourt, C., Stiell, IG., et al. Cardiac arrest care and emergency medical services in Canada. Can J Cardiol. 2004;20(11):1081–90.

17. Arntz HR, Bossaert L, Carli P, Chamberlain D, Davies M, Dellborg M, et al. The pre-hospital management of acute heart attacks. Recommendations of a Task Force of the The European Society of Cardiology and The European Resuscitation Council. Eur Heart J. Eur Heart J; 1998 Aug;19(8):1140–64.

18. Soar J, Nolan JP, Böttiger BW, Perkins GD, Lott C, Carli P, et al. European Resuscitation Council Guidelines for Resuscitation 2015: Section 3. Adult advanced life support. Resuscitation. Elsevier; 2015 Oct 1;95:100–47.

19. Digitalcommons@umaine D, Maine M, Jurson C. Incorporation of Civilian Care in Emergency Medicine: Retainment of Training and Familiarization of Resources at the University of Maine.

20. Larsen MP, Eisenberg MS, Cummins RO, Hallstrom AP. Predicting survival from out-of-hospital cardiac arrest: a graphic model. Ann Emerg Med. 1993 Nov 1;22(11):1652–8.

21. Tanaka H, Ong MEH, Siddiqui FJ, Ma MHM, Kaneko H, Lee KW, et al. Modifiable Factors Associated With Survival After Out-of-Hospital Cardiac Arrest in the Pan-Asian Resuscitation Outcomes Study. Ann Emerg Med [Internet]. Ann Emerg Med; 2018 May 1 [cited 2022 Dec 22];71(5):608-617.e15. Available from: https://pubmed.ncbi.nlm.nih.gov/28985969/

22. Wissenberg M, Lippert FK, Folke F, Weeke P, Hansen CM, Christensen EF, et al. Association of national initiatives to improve cardiac arrest management with rates of bystander intervention and patient survival after out-of-hospital cardiac arrest. JAMA. American Medical Association; 2013 Oct 2;310(13):1377–84.

23. Lancet T. Out-of-hospital cardiac arrest: a unique medical emergency. The Lancet [Internet]. 2018 [cited 2023 Feb 3];391:911. Available from: http://www.global

24. Sasson C, Haukoos JS, Ben-Youssef L, Ramirez L, Bull S, Eigel B, et al. Barriers to Calling 911 and Learning and Performing Cardiopulmonary Resuscitation for Residents of Primarily Latino, High-Risk Neighborhoods in Denver, Colorado. Ann Emerg Med. Mosby Inc.; 2015 May 1;65(5):545–552.e2.

25. Fosbøl EL, Dupre ME, Strauss B, Swanson DR, Myers B, McNally BF, et al. Association of neighborhood characteristics with incidence of out-of-hospital cardiac arrest and rates of bystander-initiated CPR: Implications for community-based education intervention. Resuscitation. Elsevier Ireland Ltd; 2014 Nov 1;85(11):1512– 7.

26. Swor RA, Jackson RE, Compton S, Domeier R, Zalenski R, Honeycutt L, et al. Cardiac arrest in private locations: Different strategies are needed to improve outcome. Resuscitation [Internet]. Elsevier Ireland Ltd; 2003 Aug 1 [cited 2023 Jun 19];58(2):171–6. Available from: http://www.resuscitationjournal.com/article/S0300957203001187/fulltext

27. Tresch DD, Thakur RK. Cardiopulmonary resuscitation in the elderly. Beneficial or an exercise in futility? Emerg Med Clin North Am. Emerg Med Clin North Am; 1998;16(3):649–63.

28. Tresch DD, Thakur R, Hoffmann RG, Brooks HL. Comparison of outcome of resuscitation of out-of-hospital cardiac arrest in persons younger and older than 70 years of age. Am J Cardiol. 1988 May 1;61(13):1120–2.

29. Alao DO, Mohammed NA, Hukan YO, Al Neyadi M, Jummani Z, Dababneh EH, et al. The epidemiology and outcomes of adult in-hospital cardiac arrest in a high-income developing country. [cited 2023 Jun 18]; Available from: 10.1016/j.resplu.2022.100220

30. Cope AR, Quinton DN, Dove AF, Sloan JP, Dave SH. Survival from cardiac arrest in the accident and emergency department. J R Soc Med. 1987;80(12):746–9.

31. Peberdy MA, Callaway CW, Neumar RW, Geocadin RG, Zimmerman JL, Donnino M, et al. Part 9: Post-cardiac arrest care: 2010 American Heart Association Guidelines for Cardiopulmonary Resuscitation and Emergency Cardiovascular Care. Vol. 122, Circulation. 2010.

32. Becker LB, Aufderheide TP, Geocadin RG, Callaway CW, Lazar RM, Donnino MW, et al. Primary outcomes for resuscitation science studies: A consensus statement from the american heart association. Circulation. 2011 Nov 8;124(19):2158–77.

